# Development and psychometric evaluation of The Index of Myalgic Encephalomyelitis Symptoms (TIMES) Part I: Rasch Analysis and Content Validity

**DOI:** 10.64898/2026.02.16.26346394

**Authors:** Mike C. Horton, Sarah F Tyson, Russell Fleming, Peter Gladwell

**Affiliations:** Psychometric Laboratory for Health Sciences, University of Leeds; School of Health Science, University of Manchester; ME Association, UK; North Bristol NHS Foundation Trust

**Author notes:** **Corresponding Author:** Prof Sarah Tyson. **Disclosure Statement:** The authors report no conflicts of interest but note that Mr Fleming is employed by the ME Association as the head of project development.

**Keywords:** Myalgic encephalomyelitis, chronic fatigue syndrome, symptoms, Rasch analysis, psychometrics, co-production

## Abstract

**Objective:** To develop and psychometrically evaluate an assessment of symptoms in myalgic encephalomyelitis/chronic fatigue syndrome (ME/CFS)

**Methods:** An initial symptom list was devised from the relevant literature with the patient and clinician advisory groups. An online survey with 85 symptom items in eight domains was completed by people with ME/CFS. Each item had two response structures (assessing symptom frequency and severity on five-point scales). Rasch analysis assessed each domain for unidimensionality, targeting, internal reliability, item fit and local dependency.

**Results:** Survey data (n=721) indicated various item anomalies and inter-item dependencies, leading to item re-formatting or removal. The frequency and severity-based responses broadly replicated each other, and a four-point response format appeared more appropriate than a five-point response format. Following Rasch-based scale amendments, a revised version with a single four-point response format was re-administered to test the modifications. Validation data (n=354) showed the modified scale had an improved response structure and functionality across all domains, satisfying Rasch model assumptions. Additionally, domain-level super-items allowed for a summated total score along with sub-scales summarising neurological and autonomic symptoms, again satisfying Rasch model assumptions.

**Conclusions:** The Index of ME Symptoms (TIMES) and its associated sub-scales and domain scales are stable, valid assessments of symptoms in ME/CFS.

## Introduction

Myalgic encephalomyelitis (ME), also referred to as chronic fatigue syndrome (CFS) is a poorly understood, chronic multi-system condition. It is classified as a neurological disease, but immune, autonomic, metabolic, and endocrine dysfunction are widely reported^1,2^. Most recent estimates indicate prevalence is 0.6%, indicating some 400,000 individuals are affected in the UK^3^. ME/CFS is highly disabling: Less than a third of people with ME/CFS are able to work^4^, a quarter are severely or very severely affected, leaving them housebound, or even bedbound^5^, and quality of life for people with ME/ CFS is lower than many other disabling chronic conditions^6^. The economic cost of ME/ CFS in the U.K. is estimated to be approximately £3.3 billion/year^7,8^.

The aetiology of ME/CFS is unclear and diagnosis is based on history and fulfilment of symptom-based diagnostic criteria. Many diagnostic criteria have been proposed, all of which are based on clinical academics’ experience^9,10^. Symptoms are categorised into domains the content of which depends on the diagnostic criteria studied, but the symptom lists and categorisations have not been psychometrically validated to establish construct validity (namely response category functioning; local dependency/ redundancy, scale targeting and item bias). Nor has face and content validity been formally evaluated.

An exception is the DePaul Symptom Questionnaire (DSQ), a self-report measure of ME/CFS symptoms. The original version has 99 questions covering personal characteristics, history and symptoms^11^. Fifty-four symptoms are included which focus on the domains specified in one of the most widely used diagnostic criteria, the Clinical Canadian Criteria (CCC)^12^. The user is asked about the frequency and severity of each symptom over the previous six months on 5-point (0-4) Likert scales. These scores are then multiplied by 25 to create 100-point ‘intensity’ scale for each symptom. The ‘intensity’ is then averaged to create a composite symptom score. The original questionnaire has been modified over time such that original, expanded, brief, and paediatric versions are now available^13^. Furthermore, a subset of questions regarding post-exertional malaise (PEM, a cardinal symptom of ME/CFS) have been extracted to produce five and ten item PEM specific questionnaires^14,15^.

As part of a project to co-produce a clinical assessment toolkit for ME/CFS with people with ME/CFS and clinicians working in NHS specialist ME/CFS services, we reviewed the DSQ for inclusion in the toolkit as a measure of symptomology. Several limitations were identified by our ME/CFS and clinicians’ advisory groups. They noted overlap/repetition between some items; use of medical jargon and ‘American English’ that was difficult to understand in some instances. They also found references to exercise and exertion problematic (as their condition meant they could not exercise or exert themselves), and having questions about both severity and frequency of symptoms did not make sense in all items and made the questionnaire very lengthy. Finally, the timescale over which symptoms were assessed (the previous six months) did not reflect the diagnostic criteria used in the UK (previous three months)^16^. Thus, we developed and evaluated a new assessment of ME/CFS symptoms focussing on the information needed for clinical assessment rather than diagnosis. This new assessment was called The Index of ME Symptoms (TIMES).

Here we present the work to develop the TIMES and evaluate the construct and content validity. Other aspects of validity, test-retest reliability and minimal detectable difference using classic theory testing are reported in a companion paper (Part 2).

## Method

The TIMES was co-produced with people with ME/CFS and clinicians working in NHS specialist ME/CFS services. An ME/CFS advisory group was convened from volunteers following publicity in the UK’s ME Association’s newsletter. Volunteers were purposively sampled to ensure men and women, and a wide range of age, geographical location, duration, and severity of ME/CFS were represented. Members of the clinical advisory group were drawn from volunteers from the British Association of Clinicians in ME (BACME), purposively sampled to ensure that a range of professions, experience working with ME and types of ME/CFS Service were represented. Both groups contributed to all stages of the project, particularly development of the assessment.

An initial prototype list of symptoms was devised from relevant literature and reviewed and revised by the advisory groups to remove any duplication, clarify descriptions and suggest additional symptoms. The symptoms were grouped into broad categories and converted into a questionnaire using five-point (0-4) Likert scales of the frequency and severity of symptoms. As the aim was to assess ‘current’ symptomology, the time over which participants scored their symptoms was *“the previous month”*.

The following descriptors were used for the scales:

- Frequency (How often have you experienced the symptom in the previous month?):

`o` I do not have this symptom
`o` Occasionally (now and then)
`o` Regularly (about half the time)
`o` Most of the time
`o` All of the time
- Severity (How severe has this symptom been in the previous month?):

`o` I do not have this symptom
`o` Mild symptoms (able to carry on with activities)
`o` Moderate symptoms (Interfering with some activities)
`o` Severe symptoms (interfering with most/ all activities)
`o` Very severe symptoms (unable carry out activities).

The resultant questionnaire included 85 symptoms in eight ‘domains’: fatigue; sleep problems; cognitive symptoms; pain and musculoskeletal symptoms; neurological symptoms; gastrointestinal symptoms; cardiovascular and respiratory symptoms; immune system symptoms.

An online version of the questionnaire was constructed using the Qualtrics survey tool. People who had been diagnosed with ME/CFS in the UK were recruited via publicity in the UK’s ME Association social media and publicity channels, ME/CFS support groups, and the 25% ME group (a charity supporting people with severe ME/CFS). They were invited to complete the questionnaire online and add any feedback about their symptomology, or the questionnaire and how it could be improved. Alternative methods of completion such as phone or video conference, or paper copies were offered for those who were unable to do so online, e.g. people with severe ME/CFS and potential participants were encouraged to contact the lead author to discuss any accommodations.

Two cohorts were recruited using the methods described above. Following initial data collection and analysis (TIMES1), a further cohort (TIMES2) were recruited who completed the revised version and the analysis repeated.

## Analysis

The Rasch Model was used for analysis^17^ and the COSMIN Design Checklist for Patient-Reported Outcome Measurement Instruments was followed to structure the analysis and reporting^18^. The Rasch Model is a unidimensional measurement model that satisfies the assumptions of fundamental measurement^19,20^, therefore providing a template against which measurement instruments can be tested. Where the intention is to sum a set of items (questions) into a single total score that represents the underlying trait, Rasch Measurement Theory (RMT) provides a unified framework for assessing several aspects of internal construct validity, where it can highlight measurement anomalies within an item set.

Rasch analysis of the data was completed with RUMM2030 software^21^, using the partial credit model^22^. The analysis process was carried out separately for each of the eight conceptually derived domains: Fatigue; Sleep; Cognitive; Musculoskeletal and pain; Neurological; Gastrointestinal; Cardiorespiratory; Immune system. The Rasch analytic process included several standard tests of fit, covering both the scale and item-levels as follows for both the frequency-based and the severity-based response structures.

- All items were assessed individually for fit to the Rasch model within the subscale item set, confirming that each item contributes to the underlying construct. Misfit was indicated where items were significant at a Bonferroni-adjusted chi-square p-value or when standardised (z-score) fit-residuals fall outside ±2.5 ^23,24^.
- Unidimensionality was evaluated by a series of t-tests^25^ with multidimensionality indicated if independent subsets of items delivered significantly different person estimates, and the lower bound 95% CI percentage of significantly different t-tests was >5%.
- Response category functioning determined whether the items’ response structure was operating in the way intended, where a functional 0-4 response category structure for each item would be indicated by sequential response thresholds (the crossover points between subsequent response categories) on the underlying logit scale^26^.
- Tests of local dependency (LD) determined whether any items in the domain were more closely related than is explained by the underlying construct, which can indicate a structural overlap or redundancy within the item set. Local dependency was indicated using a residual correlation (Q3 value) criterion cut point of 0.2 above average residual correlation^27^.
- Item bias was assessed through uniform and non-uniform differential item functioning (DIF) testing by sex and age group, with significant DIF indicated at a Bonferroni-adjusted ANOVA p-value.

Scale targeting was assessed graphically through the relative distribution of item and person locations. Reliability indices were taken as the person separation index (PSI) and Cronbach’s alpha values^23^.

Once fit to the Rasch model has been demonstrated, it is possible to transform the ordinal raw scores of the scales into interval-level data^28^. Thus, conversion charts were created for each separate domain with all scores converted to 0-100 scales to allow for easier interpretation. Additionally, conversion charts were also created for the TIMES total score and the neurological and autonomic dysfunction sub-scales. These conversion charts provide the means to take advantage of continuous data properties in new samples and allow for parametric analyses to be performed using TIMES data.

Content validity was examined using thematic content analysis of participants’ feedback. As the information sought was specific to the feasibility and acceptability of the questionnaire, a deductive approach was used. Initially participants’ feedback regarding the questionnaire was categorised into ‘survey format’, ‘item format’, ‘response options’, ‘wording of items’ and ‘suggested additions’. These were sub-categorised iteratively and a further category of ‘positive feedback’ was added during the analysis. The feedback data were initially independently categorised by two of the authors who then came together to compile and integrate their findings. The results of the analysis were discussed with the other research team and advisory group members by way of external member checking to ensure trustworthiness/external validity.

## Results

The two cohorts of participants had very similar characteristics (Table 1). Reflecting the demographics of ME/CFS, they were predominantly middle-aged women with moderately severe ME/CFS and extensive lived experience.

**Table 1.** Characteristics of the recruited cohorts.

|  | **TIMES 1 (n=701)** | **TIMES 2 (n354)** |
| --- | --- | --- |
| Gender (m/f/other) | 116 (16%)<br>572 (82%)<br>13 (2%) | 58 (16.4%)<br>295 (83%)<br>1 (0.3%) |
| Age (Mean, sd) | 53.7 (13.5) | 56.3 (13.6) |
| Duration (mean, sd) | 14.7 (12.2) | 16.2 (12.2) |
| Severity | Mean (sd) 2.85 (0.89)<br>L1: 41 (5.8%)<br>L2: 194 (27.7%)<br>L3: 310 (44.2%)<br>L4: 138 (19.7%)<br>L5: 18 (2.6%) | 2.9 (0.95)<br>L1: 23 (6.5%)<br>L2: 100 (28.2%)<br>L3: 132(37%)<br>L4: 89(25%)<br>L5: 10 (2.8%) |
| **Location** |  |  |
| England | 606 (86%) | 309 (87%) |
| Wales | 25 (3.6%) | 1 (3.4%) |
| Scotland | 62 (8.8) | 30 (8.5%) |
| Northern Ireland | 8 (1.1%) | 0 |

*L1= Level 1, very mild ME/CFS; L2 = Level 2, mild ME/CFS; L3= level 3, moderately severe ME/CFS; L4= level 4 severe ME/CFS; L5= level 5, very severe ME/CFS.*

The process to validate the TIMES and the main changes made are summarised in Figure 1.

**Figure 1.**
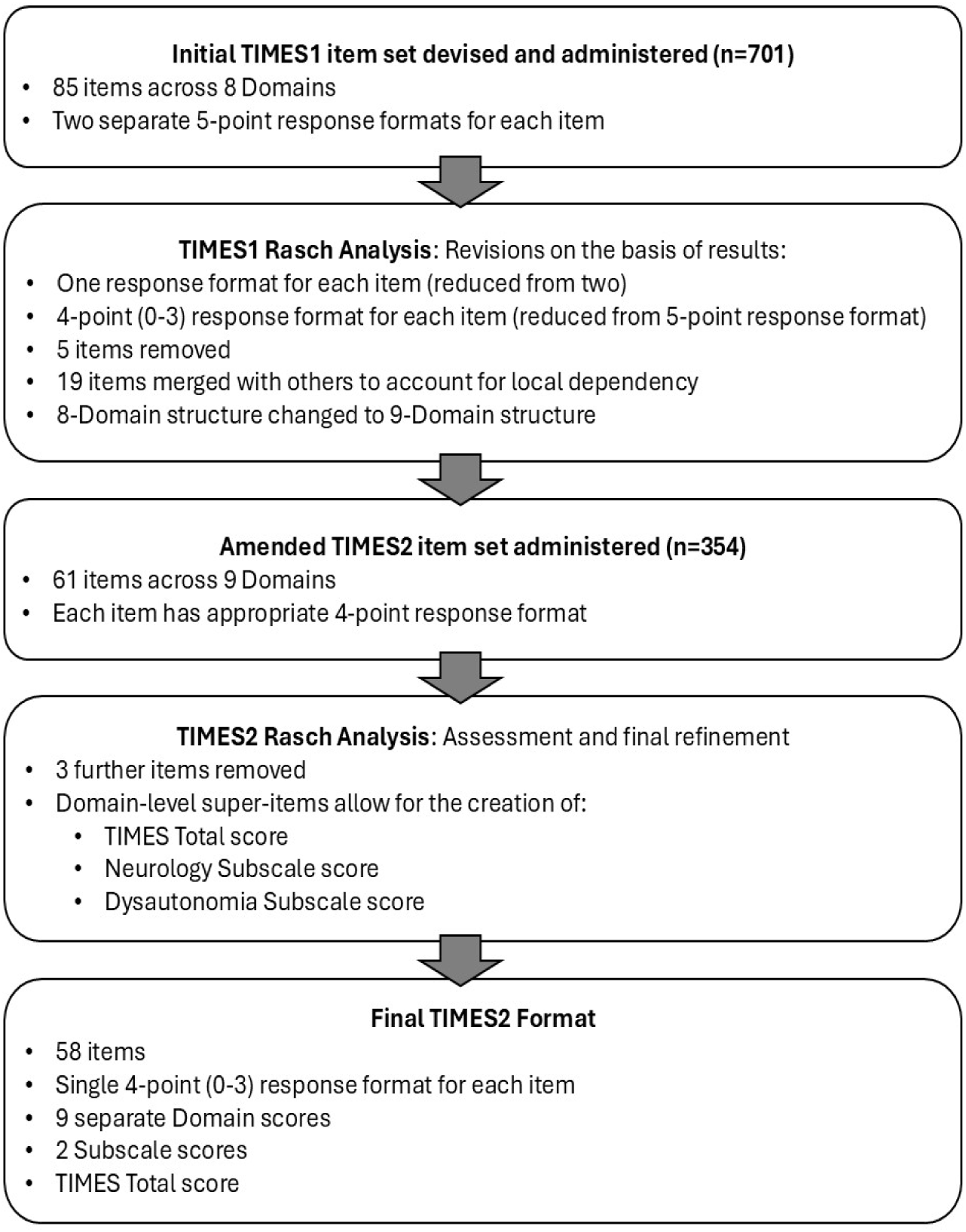
The process of development and validation of The Index of ME Symptoms (TIMES)

The results of the TIMES1 analyses are summarised in Table 2.

**Table 2.** Summary of TIMES1 Rasch Analyses across both response formats of the eight TIMES1 domains.

| **Domain** | **No. of items** | **Response category** | **Disordered items (n)** | **Misfit items (n) \*** | **Local Dependency (n)** | **Unidimensionality** | **Person Separation Index** |
| --- | --- | --- | --- | --- | --- | --- | --- |
| Fatigue | 8 | Frequency | 1 | 3 | 5/28 | 9.36% | 0.87 |
|  |  | Severity | 7 | 4 | 5/28 | 8.20% | 0.88 |
| Sleep | 10 | Frequency | 8 | 4 | 6/45 | 8.07% | 0.74 |
|  |  | Severity | 9 | 6 | 6/45 | 12.70% | 0.85 |
| Cognitive | 15 | Frequency | 5 | 8 | 9/105 | 8.85% | 0.93 |
|  |  | Severity | 7 | 8 | 11/105 | 11.62% | 0.94 |
| Pain/MSK | 15 | Frequency | 10 | 2 | 6/105 | 7.70% | 0.86 |
|  |  | Severity | 12 | 5 | 5/105 | 7.70% | 0.89 |
| Neurological | 12 | Frequency | 9 | 2 | 3/66 | 2.90% | 0.8 |
|  |  | Severity | 8 | 5 | 3/66 | 5.42% | 0.84 |
| GI | 12 | Frequency | 9 | 4 | 3/66 | 3.25% | 0.69 |
|  |  | Severity | 9 | 3 | 1/66 | 3.87% | 0.73 |
| CR | 8 | Frequency | 6 | 0 | 2/21 | 3.34% | 0.66 |
|  |  | Severity | 5 | 0 | 1/21 | 3.99% | 0.74 |
| Immune | 4 | Frequency | 3 | 0 | 0/6 | 0.53% | 0.33 |
|  |  | Severity | 1 | 1 | 0/6 | 1.41% | 0.47 |

\* = Number of items displaying either a Chi-square or Fit Residual misfit. MSK = musculoskeletal, GI = gastrointestinal, CR= cardiorespiratory

It was observed that the frequency-based and the severity-based items broadly replicated each other, in terms of the item location ordering and the items identified as misfitting or displaying local dependency. However, it was also noted that in terms of the response structure, some of the domains displayed a definite preference towards one format over the other. For example, the response structure of the Fatigue domain appeared to operate much better with frequency-based responses than with severity-based responses. Upon review of the item content, this aligned with how people would respond to the items, with the frequency-based responses appearing to be more appropriate within this domain.

Due to the replication of results, each domain was reduced to one response format, and the most appropriate response format was selected for each domain, based on the empirical response ordering and the item content. The severity-based response structure was most appropriate for all domains except Fatigue and Cognitive, where frequency-based responses were chosen.

Alongside the selection of a single response format, it was apparent that for most items, the response structure wasn’t working as intended, and disordered response thresholds were observed. This was consistently due to the non-emergence of the second response category across both the frequency-based and severity-based formats. A generic post-hoc rescoring was therefore implemented across all items, where the second and third response options were merged. Thus, the response ‘Occasionally (now and then)’ was merged with ‘Regularly (about half the time)’, and ‘Mild symptoms (able to carry on with activities)’ was merged with ‘Moderate symptoms (Interfering with some activities)’. This vastly reduced the number of items displaying disordered thresholds.

Within the analysis of all domains, several individually misfitting items were observed alongside many items displaying local dependency (see Table 2). Based on these results and a content-based analysis of the affected items, an iterative process of domain amendment was carried out. Nineteen were merged into other items to account for the observed local dependency. For example, the item *‘Difficulty getting started on tasks’* was shown to be locally dependent with the item *‘Difficulty finishing or completing tasks’*, so in the TIMES2 item set these items were combined into a single item stating *‘Difficulty starting and/or finishing tasks’*.

The item misfit patterns and local dependency clustering also led to a conceptual restructuring of the domains, where a further domain (‘Cranial Nerves’) was introduced, and some items moved between domains. This restructuring resulted in the items representing nine domains, instead of the original eight. Where items still remained as anomalies following all post-hoc amendments and domain restructuring, they were completely removed from the item set (n=5).

During analysis of TIMES2 (Figure 1), no further post-hoc modelling adjustments were made to the item set, but three further items were removed due to excessive misfit: The ‘wired but tired’ item was removed from the Cranial Nerves domain; the ‘bladder’ item was removed from the Gastrointestinal domain; and the ‘alcohol intolerance’ item was removed from the Immune System domain. All other items displayed Rasch model fit within their domains, although some minor anomalies remained. This included some locally dependent items, some items displaying differential item functioning, and one item displaying disordered response thresholds. However it was felt that the clinical and psychometric information provided by these items outweighed any potential issues impacting the total domain scores and they were retained.

The final TIMES2 item set consists of 58 items across the nine previously stated domains (fatigue, cognition, pain, motor-sensory, sleep, cranial nerves, cardio-respiratory, gastro-intestinal, and immune system) and the Rasch model fit and psychometric properties of the domains are summarised in Table 3. Further information relating to the individual item fit of the domains can be found in Supplementary Tables 1.

**Table 3.** Summary of the psychometric properties of the nine individual domains of the TIMES2 scale.

|  | **Fatigue** | **Cognitive** | **Pain** | **Motor-sensory** | **Cranial Nerves** | **GI** | **CR** | **Sleep** | **Immune** | **Target Values** |
| --- | --- | --- | --- | --- | --- | --- | --- | --- | --- | --- |
| total n - (extremes) = valid n | **354 - (45) = 309** | **354 - (1) = 353** | **354 - (9) = 345** | **354 - (5) = 349** | **354 - (8) = 346** | **354 - (8) = 346** | **354 - (2) = 352** | **354 - (2) = 352** | **354 - (32) = 322** |  |
| Number of items | **4** | **9** | **6** | **7** | **6** | **7** | **9** | **6** | **4** |  |
| Overall Scale Fit (Chi-square p) | **0.19** | **0.07** | **0.19** | **0.95** | **0.02** | **0.15** | **0.05** | **0.12** | **0.43** | p>0.01 |
| Individual Item Fit Residuals (n items out of range) | none | none | none | none | none | none | none | none | none | within +/- 2.5 |
| Individual Item Fit (Chi-square p) (n items out of range) | none | none | none | none | none | none | none | none | none | p>0.05* |
| Person Fit Residuals (% outside +/-3.0) | 0.00% | 2.26% | 1.13% | 5.37% | 1.98% | 0.56% | 0.85% | 3.11% | 11.30% | within +/- 3.0 |
| Unidimensionality | 0.97% | 7.37% (5.1%) | 4.93% | 8.31% (6.0%) | 3.47% | 4.91% | 9.09% (6.8%) | 6.82% (4.5%) | 2.48% | % sig t-tests <5% (Lower 95% CI) |
| Local Dependency | none | 2/36 pairwise significant | 1/15 pairwise significant | 1/21 pairwise significant | none | 1/21 pairwise significant | 4/36 pairwise significant | 1/15 pairwise significant | none | <0.2 above average Q3 |
| Targeting | right skew | good alignment | slight left skew | slight left skew | slight left skew | left skew | slight left skew | good alignment | left skew | Aligned |
| % sample at floor (min score) | 0.0% | 0.0% | 2.5% | 1.4% | 2.3% | 2.3% | 0.6% | 0.0% | 9.0% |  |
| % sample at ceiling (max score) | 12.7% | 0.3% | 0.0% | 0.0% | 0.0% | 0.0% | 0.0% | 0.6% | 0.0% |  |
| % sample at floor and ceiling | 12.7% | 0.3% | 2.5% | 1.4% | 2.3% | 2.3% | 0.6% | 0.6% | 9.0% | <15% |
| (combined) |  |  |  |  |  |  |  |  |  |  |
| Person Separation Index (PSI) | 0.74 | 0.9 | 0.78 | 0.83 | 0.68 | 0.74 | 0.81 | 0.76 | 0.67 | >0.85 |
| Cronbach's Alpha | 0.82 | 0.91 | 0.81 | 0.86 | 0.73 | 0.79 | 0.84 | 0.78 | 0.77 | >0.85 |
| Response category threshold ordering | all ordered | all ordered | all ordered | all ordered | all ordered | all ordered | 1/9 disordered | all ordered | all ordered | All ordered |
| Differential Item Function by Sex | None | None | 2/6 items | None | Borderline 1 item | None | Borderline 1 item | None | None | p>0.05* |
| Differential Item Function by Age Group | Borderline 1 item | None | None | None | 1/6 items | 1/7 items | None | None | None | p>0.05* |

\* (Bonferroni adjusted) GI = gastro-intestinal CR = cardio-respiratory

In addition to the nine-domain score profile which can be established for each person, it is also useful to summarise a person’s overall level of symptom burden with a single total score. Thus, the items within each domain were combined to create domain-level ‘super-items’, resulting in a 9-item scale where each item represents a domain. This was named the TIMES-Total scale. The results of the analyses to create the TIMES-Total scale are presented in Table 4 under the name ‘DOMAIN Items’, and further information relating to the individual item fit of the domain-items can be found in Supplementary Tables 2. It is acknowledged that there are some sources of misfit within this domain-item structure, and a degree of multidimensionality is present when the items are considered in this way. Nevertheless, given the psychometric strength of the underlying subscale scores and the low number of issues observed, it was felt that the relative degree of misfit was a fair trade-off for clinical benefit of having total scores to summarise a person’s level of symptom burden.

**Table 4.** Summary of the psychometric properties of the Neurology and Dysautonomia sub-scales, and the TIMES2 total scale, using domain-level super-items.

|  | **Neurology**<br>**Subscale** | **Dysautonomia**<br>**Subscale** | **DOMAIN**<br>**Items** | **Target Values** |
| --- | --- | --- | --- | --- |
| total n - (extremes) = valid n | **354 - (0) = 354** | **354 - (0) = 354** | **354 - (0) = 354** |  |
| Number of items | **3** | **5** | **9** |  |
| Overall Scale Fit (Chi-square p) | **0.008** | **0.43** | **<0.001** | p>0.01 |
| Individual Item Fit Residuals (n items out of range) | 1 (borderline) | 1 | 3 | within +/-2.5 |
| Individual Item Fit (Chi-square p) (n items out of range) | none | none | 1 | p>0.05 (Bonferroni adjusted) |
| Person Fit Residuals (% outside +/-3.0) | 2.26% | 1.69% | 1.98% | within +/-3.0 |
| Unidimensionality | 9.32% lower CI = 7.1% | 7.06% lower CI = 4.8% | 11.86%(lower CI = 9.6% | % of significant t-tests <5% |
| Local Dependency | 1/3 pairwise significant | none | 2/36 pairwise significant | <0.2 above average Q3 |
| Targeting | good alignment | very slight left skew | good alignment | aligned |
| % sample at floor (min score) | 0.0% | 0.0% | 0.0% |  |
| % sample at ceiling (max score) | 0.0% | 0.0% | 0.0% |  |
| % sample at floor and ceiling (combined) | 0.0% | 0.0% | 0.0% | <15% |
| Person Separation Index (PSI) | 0.82 | 0.87 | 0.93 | >0.85 |
| Cronbach's Alpha | 0.8 | 0.87 | 0.92 | >0.85 |
| Response category threshold ordering | all ordered | all ordered | all ordered | All ordered |
| Differential Item Functioning by Sex | 2/3 items | None | None | p>0.05 (Bonferroni adjusted) |
| Differential Item Functioning by Age Group | None | 1/5 items | 1/9 items | p>0.05 (Bonferroni adjusted) |

The domain-level super-items (i.e. TIMES Total scale) were also partitioned into two further subscales – a ‘Neurology’ subscale comprising the Cognitive, Pain and Motor-sensory domains; and a ‘Dysautonomia’ subscale comprising the Sleep, Cranial Nerves, Gastrointestinal, Cardiorespiratory and Immune system domains. The results of these subscale analyses are also presented in Table 4, and further information relating to the individual item fit of these subscales can be found in Supplementary Tables 2. The TIMES portfolio of measures is summarised in Figure 2

**Figure 2.**
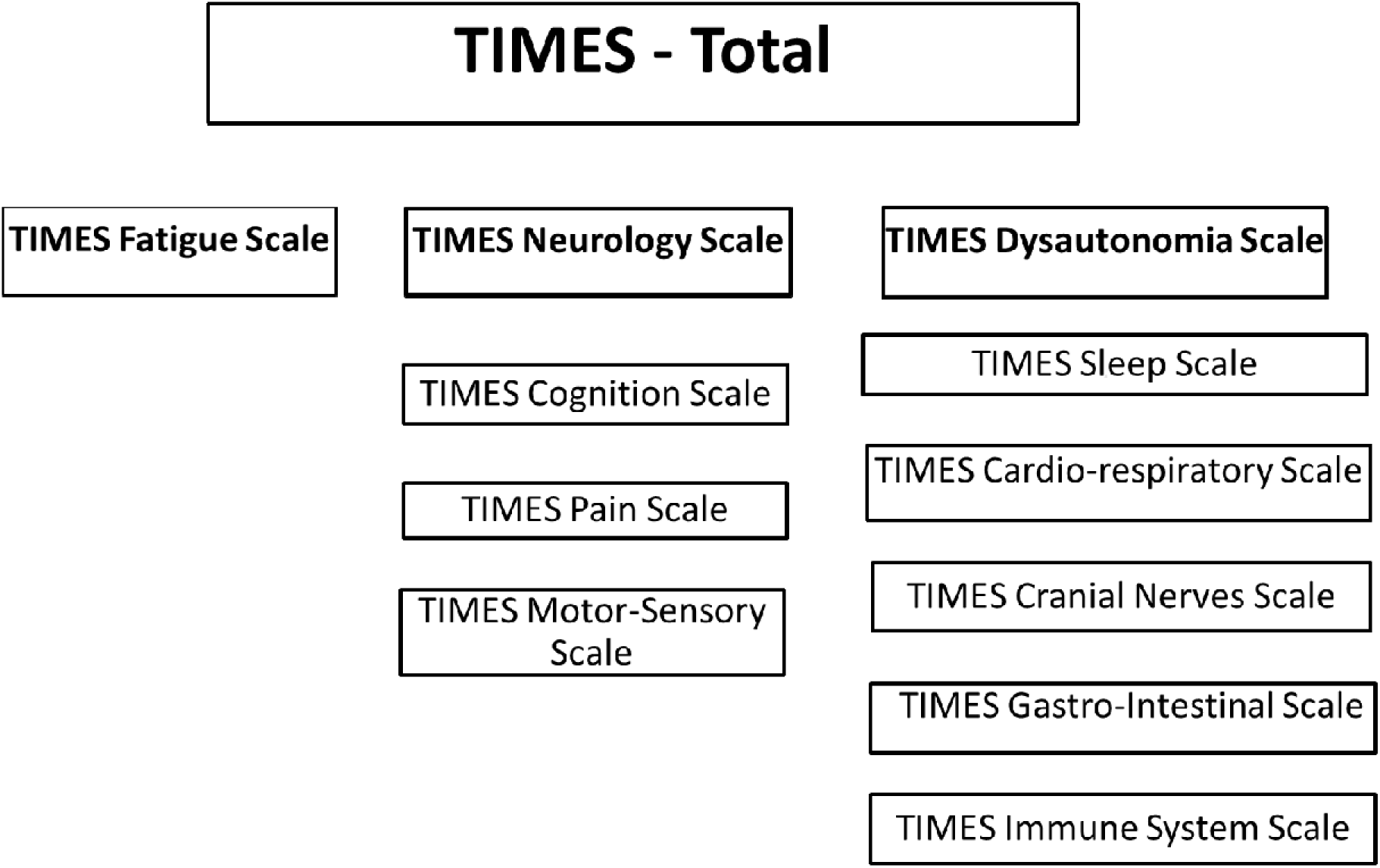
The TIMES Portfolio of Assessments.

The conversion tables to transform the ordinal score data to interval are presented in Supplementary Table 3. It should be noted that these conversion charts are only valid in the case of complete data.

## Content validity

Overall feedback from participants was positive about the TIMES, with many expressing gratitude that their lived experience had been captured:

*Really excellent questions and options to answer. I can’t believe (after much medical gaslighting) how much this has described my symptoms and new life experience. Thank you! (PTID556)*

Many participants explained that they had co-existing conditions with symptoms which overlapped with ME/CFS. To address this, we resolved to formulate and evaluate a co-morbidities checklist so people could record any other conditions which they felt impacted on their ME/CFS symptomology. This will be published in due course.

A few participants suggested further symptoms that could be added, however there was no consistency in the symptoms suggested. Some reported that they found it difficult to generalise about their symptoms over the previous month because they fluctuated so much. To accommodate these preferences, free text boxes were added to each domain so people could add further detail if they wished.

There were some suggestions about how to make the questionnaire easier to complete. To address this, the following details were added:

- Detail in the introduction re: what was involved in completing, how long it was likely to take and that it was fine to have assistance to complete.
- Automatic saving; a back button (to check answers); a ‘% Completed’ tally (to enable pacing).
- Instruction that answers should reflect what was ‘normal for them’, rather than compared to healthy people, before they became ill or other people with ME/CFS.
- Guidance about how respondents should answer individual items if they were unable to undertake relevant activities. For example, those who are artificially feed should answer ‘very severe’ to the question about difficulty eating and drinking.

## Discussion

The results of this study have shown that the TIMES, the subscales and the domain scales are robust, valid measures of symptomology in ME/CFS. The TIMES and the sub-scales give an overview of how people experience their symptoms, while the domain scales can be used as stand-alone assessments of more specific aspects of ME/CFS. Co-production with people with ME/CFS and clinicians working in NHS specialist ME/CFS services has ensured the items are relevant and important to both people with ME/CFS and clinicians, and it is feasible and acceptable to complete.

This study presents the complete cycle of item set development, psychometric analysis, empirical modification, and psychometric validation of the modified scale in a new cohort. It demonstrates the improvements that can be made in the psychometric performance of an instrument by basing modifications on empirical evidence. It is rare to see scale development studies demonstrating this complete analytical loop, therefore providing evidence of the depth and diligence of the scale development approach. The final version of the TIMES is shown in Supplementary File 4 and is freely available for others to use. A digital version is available via the Autono-ME app (<u>The ME Association App</u>) and ‘clinic-ready’ hard copies can be downloaded from <u>The ME Association Clinical Assessment Toolkit (MEA-CAT) - The ME</u> <u>Association</u>

The use of Rasch analysis has demonstrated that the two-stage response option (asking about both frequency and severity of symptoms) largely duplicated answers i.e. people tended to give the same answer to both questions. A two-question response was initially chosen as it was anticipated that infrequent, severe symptoms, or frequent, mild symptoms would occur. However, this was rarely the case, rather symptom severity tended to increase with frequency. Additionally, item fit results tended to be replicated across both response formats. For example, if an item appeared to be a misfit anomaly or to display local dependency for the ‘frequency’ response format, the same findings occurred in the ‘severity’ response format of the item. By reducing the response options to a single format, the demands of completion were halved, as well as avoiding score inflation. Rasch analysis also provided empirical evidence that a four-item response option was more effective than five, which further reduced completion demands. The removal and merging of redundant or dependent items also reduced completion demands, whilst also improving the psychometric properties of the item set.

The use of Rasch methodology focussed on the quality of measurement for each of the domains. This set of scores therefore creates a detailed disease profile for each individual, which is clinically useful and could provide a basis for future phenotyping studies as well as development protocols to manage the symptoms. Alongside the disease profile, the scale also provides higher level scores for ‘Neurology’ and ‘Dysautonomia’ subscales, and when a single total score is also required to represent an individual’s location on the underlying trait, an overall TIMES total score is also available. The use of Rasch methodology allows for interval-level transformations of each of these scores, either at the level of the domain, subscale or total scale which enables future analyses using psychometric statistics.

The use of Rasch analysis has also revealed important insights into ME/CFS symptomology. The detailed, empirically derived exploration of how items/symptoms clustered together resulted in a framework conceptualising ME/CFS symptoms. The most relevant previous study is Brown and Jason’s 2014 paper^29^ which validated the DSQ. This used exploratory factor analysis and reported three factors (Neuroendocrine, Autonomic, and Immune Symptoms; Neurological/Cognitive Dysfunction; Post-Exertional Malaise). These broadly concur with the TIMES’ dysautonomia, neurological and fatigue sub-scales. Further confirmation is provided in Conroy et al’s (2022)^30^ factor analysis of a very large international sample of people with ME/CFS using the DSQ. It incorporated a review of other factor analysis studies against the numerous diagnostic criteria for ME/CFS, showing the overlap and inconsistencies between them. It concluded on six symptom categories (cognition; dysautonomia (sub-divided into orthostatic intolerance and immune dysfunction); post-exertional malaise, sleep; circulatory/ neuroendocrine dysfunction, and gastro-intestinal distress). These broadly equate to the TIMES scales, with the exception of pain, cranial nerves and motor-sensory symptoms which were found in the TIMES. These symptom categories were present in the DSQ but were dropped during Conroy et al.’s analysis^30^ due to ‘low factor loading’. To our knowledge, these DSQ symptom categories have not been evaluated as stand-alone scales.

## Limitations

The strengths of this study lie in the large, representative sample, the robust co-production with people with ME/CFS and clinicians working in NHS specialist ME/CFS services. The number of people with severe/very severe ME/CFS recruited is also a strength. It is estimated that 25% of people with Me/CFS are severely or very severely affected^5^. This is reflected in the TIMES cohorts and is higher than other large studies using similar recruitment methods, such as DeCodeME^31^. We think the accommodations offered to maximise accessibility enabled a relatively high proportion of severely affected people to participate. These included alternative ways to complete the surveys (paper, phone, video or proxy); a survey tool that automatically saved responses so it could be completed in phases; using a dyslexia friendly style guide to maximise accessibility, and encouraging discussion with researchers and (potential) participants about their needs and further accommodations.

A further strength is the thoroughness with which the Rasch analysis was completed. It is unusual to see the complete cycle of scale development, analysis, empirical modification, and psychometric validation of the modified scale in a new cohort, and the conversion tables to enable parametric analyses are rarely presented.

Possible limitations lie in the representativeness of the sample. The literature regarding the demographics of ME./CFS are sparse, so it is not possible to compare our sample with an authoritative source of epidemiological data. However, our demographics broadly reflect other large ME/CFS studies with convenience samples^31–34^: predominantly middle-aged women with moderately severe illness and extensive lived experience. Thus, we are confident data are reasonably representative of the ME/ CFS community. However, our recruitment methods would have biased towards English speakers who are active on social media and involved with ME/CFS organisations. We recruited people with a ‘formal diagnosis’ of ME/CFS, but we did not confirm the diagnosis via health records. This was a deliberate choice because the recording of ME/CFS in both primary and secondary care is notoriously inaccurate, incomplete and inconsistent^33,35^. However, it does mean that we may have recruited some people who would prove to have other diagnoses if thoroughly assessed or accurately recorded. Further research is needed to better understand the demographics of ME/CFS and to accurately diagnose and record it. Finally, we specifically limited recruitment to adults and so the TIMES would need to be validated for children and young people with ME/CFS before implementation with this population.

## Supporting information

Supplemental File 1. Indiviv Item Fit Domains

Supplementary File 2 Individ Item Fit Subscales

Supplementary file 3. Rasch ordinal to interval conversion tables

Supplementary file 4 TIMES final version

## Data Availability

All data produced in the present study are available upon reasonable request to the authors

## Acknowledgements

The authors would like to thank the members of the advisory groups for their unstinting support and the invaluable knowledge and insights they provided.

