## Supplemental File 1. Indiviv Item Fit Domains for "Development and psychometric evaluation of The Index of Myalgic Encephalomyelitis Symptoms (TIMES) Part I: Rasch Analysis and Content Validity"

**Supplementary File 1 – Rasch Analysis Final Individual Item Fit Tables for each TIMES Domain**

Any highlighted (coloured) values fall outside of fit criteria stated in Methods section

### Fatigue Domain Individual Item Fit Table

| | | | | | | Local Dependency | | | |
|---|---|---|---|---|---|---|---|---|---|
| Item Code | Statement | Location | SE | Fit Residual | Chi-Square P | FA1 | FA2 | FA3 | FA4 |
| FA1 | Exhaustion | -0.374 | 0.108 | -1.229 | 0.090 | | | | |
| FA2 | Loss of stamina | 0.058 | 0.104 | -0.947 | 0.194 | -0.157 | | | |
| FA3 | Brain fog | 0.243 | 0.108 | 1.566 | 0.205 | -0.348 | -0.377 | | |
| FA4 | PEM | 0.072 | 0.103 | 0.153 | 0.944 | -0.322 | -0.336 | -0.379 | |
| | | | | | Average residual correlation | -0.320 | | | |
| | | | | | LD criterion (ave + 0.2) | -0.120 | | | |

### Cognitive Domain Individual Item Fit Table

| | | | | | | Local Dependency | | | | | | | | |
|---|---|---|---|---|---|---|---|---|---|---|---|---|---|---|
| Item Code | Statement | Location | SE | Fit Residual | Chi-Square P | CO1 | CO2 | CO3 | CO4 | CO5 | CO6 | CO7 | CO8 | CO9 |
| CO1 | Memory/ concentration | -1.062 | 0.102 | 0.696 | 0.864 | | | | | | | | | |
| CO2 | Slow thought | -0.125 | 0.101 | -1.361 | 0.451 | 0.195 | | | | | | | | |
| CO3 | Starting /finishing tasks | -0.368 | 0.095 | -1.201 | 0.092 | -0.197 | -0.22 | | | | | | | |
| CO4 | Decision-making/ problem solving | 0.187 | 0.095 | -1.95 | 0.071 | -0.168 | -0.029 | -0.024 | | | | | | |
| CO5 | Difficulty getting organised | 0.665 | 0.092 | -0.215 | 0.610 | -0.272 | -0.287 | 0.094 | 0.023 | | | | | |
| CO6 | Multi-tasking | -0.157 | 0.087 | 1.728 | 0.060 | -0.286 | -0.124 | -0.008 | -0.148 | -0.009 | | | | |
| CO7 | Understanding information | -0.065 | 0.097 | -1.723 | 0.194 | -0.060 | -0.121 | -0.133 | -0.12 | -0.234 | -0.159 | | | |
| CO8 | Communication | -0.033 | 0.102 | 0.656 | 0.284 | 0.066 | -0.029 | -0.316 | -0.181 | -0.235 | -0.273 | 0.009 | | |
| CO9 | Reading/ writing | 0.958 | 0.093 | 2.004 | 0.304 | -0.161 | -0.206 | -0.175 | -0.229 | -0.141 | -0.2 | -0.096 | 0.078 | |
| | | | Average residual correlation | | | -0.122 | | | | | | | | |
| | | | LD criterion (ave + 0.2) | | | 0.078 | | | | | | | | |

### Motor-Sensory Domain Individual Item Fit Table

| | | | | | | Local Dependency | | | | | | |
|---|---|---|---|---|---|---|---|---|---|---|---|---|
| Item Code | Statement | Location | SE | Fit Residual | Chi-Square P | MS1 | MS2 | MS3 | MS4 | MS5 | MS6 | MS7 |
| MS1 | Muscle tightness | -0.427 | 0.086 | 0.018 | 0.310 | | | | | | | |
| MS2 | Muscle cramps | 0.2 | 0.091 | -0.779 | 0.796 | 0.079 | | | | | | |
| MS3 | Tremors/ shakiness | 0.721 | 0.096 | -0.374 | 0.777 | -0.239 | -0.18 | | | | | |
| MS4 | Slow movement | -0.567 | 0.09 | 1.103 | 0.625 | -0.177 | -0.337 | 0.006 | | | | |
| MS5 | Clumsiness/ balance problems | -0.358 | 0.091 | -0.049 | 0.684 | -0.309 | -0.182 | -0.058 | -0.078 | | | |
| MS6 | Touch/ pressure sensitivity | 0.44 | 0.09 | 0.026 | 0.634 | -0.134 | -0.256 | -0.258 | -0.114 | -0.22 | | |
| MS7 | Numbness | -0.009 | 0.088 | -0.045 | 0.923 | -0.250 | -0.062 | -0.159 | -0.357 | -0.136 | -0.026 | |
| | | | Average residual correlation | | | -0.164 | | | | | | |
| | | | LD criterion (ave + 0.2) | | | 0.036 | | | | | | |

### Pain Domain Individual Item Fit Table

| | | | | | | Local Dependency | | | | | |
|---|---|---|---|---|---|---|---|---|---|---|---|
| Item Code | Statement | Location | SE | Fit Residual | Chi-Square P | PA1 | PA2 | PA3 | PA4 | PA5 | PA6 |
| PA1 | Musculo-skeletal Pain | -1.281 | 0.092 | -0.886 | 0.256 | | | | | | |
| PA2 | Jaw pain | 1.095 | 0.091 | -0.08 | 0.438 | -0.209 | | | | | |
| PA3 | Eye pain | 0.805 | 0.09 | -0.474 | 0.644 | -0.334 | -0.133 | | | | |
| PA4 | Neuralgia | -0.427 | 0.082 | -1.817 | 0.122 | 0.072 | -0.225 | -0.22 | | | |
| PA5 | Headaches /migraines | -0.111 | 0.085 | 2.413 | 0.096 | -0.331 | -0.17 | -0.021 | -0.325 | | |
| PA6 | Allodynia | -0.082 | 0.083 | 0.015 | 0.584 | -0.072 | -0.2 | -0.224 | -0.198 | -0.323 | |
| | | | | | Average residual correlation | -0.194 | | | | | |
| | | | | | LD criterion (ave + 0.2) | 0.006 | | | | | |

### Cardio-Respiratory Domain Individual Item Fit Table

| Item Code | Statement | Location | SE | Fit Residual | Chi-Square P | Local Dependency | | | | | | | | |
|---|---|---|---|---|---|---|---|---|---|---|---|---|---|---|
| | | | | | | CR1 | CR2 | CR3 | CR4 | CR5 | CR6 | CR7 | CR8 | CR9 |
| CR1 | Temperature intolerance/ sensitivity | -1.086 | 0.086 | -1.967 | 0.030 | | | | | | | | | |
| CR2 | Dizziness | 0.034 | 0.086 | 0.717 | 0.083 | -0.140 | | | | | | | | |
| CR3 | Palpitations | -0.087 | 0.086 | -1.081 | 0.619 | -0.198 | -0.166 | | | | | | | |
| CR4 | Chest pain | 1.182 | 0.094 | -0.362 | 0.809 | -0.208 | -0.122 | 0.032 | | | | | | |
| CR5 | Shortness of breath | 0.041 | 0.084 | 0.028 | 0.578 | -0.217 | -0.193 | 0.072 | 0.11 | | | | | |
| CR6 | Poor circulation | -0.425 | 0.079 | 0.877 | 0.522 | 0.128 | -0.262 | -0.188 | -0.103 | -0.171 | | | | |
| CR7 | Orthostatic intolerance | -0.472 | 0.079 | -0.859 | 0.599 | -0.276 | 0.096 | -0.043 | -0.17 | -0.119 | -0.225 | | | |
| CR8 | Swollen extremities | 0.648 | 0.082 | -0.072 | 0.019 | -0.145 | -0.112 | -0.169 | -0.171 | -0.165 | -0.086 | -0.075 | | |
| CR9 | Abnormal sweating | 0.164 | 0.079 | 0.907 | 0.145 | 0.099 | -0.116 | -0.193 | -0.201 | -0.206 | -0.176 | -0.232 | -0.071 | |
| | | | Average residual correlation | | | -0.122 | | | | | | | | |
| | | | LD criterion (ave + 0.2) | | | 0.078 | | | | | | | | |

### Gastro-Intestinal Domain Individual Item Fit Table

| | | | | | | Local Dependency | | | | | | |
|---|---|---|---|---|---|---|---|---|---|---|---|---|
| Item Code | Statement | Location | SE | Fit Residual | Chi-Square P | GI1 | GI2 | GI3 | GI4 | GI5 | GI6 | GI7 |
| GI1 | Nausea | 0.299 | 0.091 | 1.525 | 0.222 | | | | | | | |
| GI2 | Abdominal pain | -0.576 | 0.087 | -1.542 | 0.145 | -0.105 | | | | | | |
| GI3 | Excess flatulence | -0.199 | 0.082 | 1.105 | 0.215 | -0.237 | 0.026 | | | | | |
| GI4 | Bowel habit | -0.814 | 0.083 | -1.397 | 0.316 | -0.204 | -0.089 | -0.135 | | | | |
| GI5 | Appetite | -0.19 | 0.083 | -0.687 | 0.265 | -0.171 | -0.263 | -0.257 | -0.117 | | | |
| GI6 | Difficulty eating and drinking | 0.961 | 0.099 | 0.022 | 0.752 | -0.163 | -0.235 | -0.175 | -0.212 | -0.173 | | |
| GI7 | Too tired to eat | 0.519 | 0.094 | 0.748 | 0.272 | -0.161 | -0.299 | -0.352 | -0.249 | -0.009 | 0.159 | |
| | | | | | Average residual correlation | -0.163 | | | | | | |
| | | | | | LD criterion (ave + 0.2) | 0.037 | | | | | | |

### Sleep Domain Individual Item Fit Table

| | | | | | | Local Dependency | | | | | |
|---|---|---|---|---|---|---|---|---|---|---|---|
| Item Code | Statement | Location | SE | Fit Residual | Chi-Square P | SL1 | SL2 | SL3 | SL4 | SL5 | SL6 |
| SL1 | Altered Sleep pattern | 0.722 | 0.079 | 0.385 | 0.087 | | | | | | |
| SL2 | Napping during the day | 0.241 | 0.085 | 1.766 | 0.483 | -0.213 | | | | | |
| SL3 | Difficulty falling asleep | 0.267 | 0.085 | -0.712 | 0.421 | -0.078 | -0.278 | | | | |
| SL4 | Poor sleep maintenance | -0.305 | 0.084 | 0.498 | 0.408 | -0.121 | -0.23 | -0.048 | | | |
| SL5 | Long time to come to | -0.041 | 0.079 | -0.091 | 0.105 | -0.314 | -0.232 | -0.267 | -0.278 | | |
| SL6 | Feeling flu-like on waking | -0.885 | 0.086 | -0.783 | 0.316 | -0.324 | -0.101 | -0.252 | -0.316 | 0.091 | |
| | | | | | Average residual correlation | -0.197 | | | | | |
| | | | | | LD criterion (ave + 0.2) | 0.003 | | | | | |

### Cranial Nerves Domain Individual Item Fit Table

| Item Code | Statement | Location | SE | Fit Residual | Chi-Square P | Local Dependency CN1 | CN2 | CN3 | CN4 | CN5 | CN6 |
|---|---|---|---|---|---|---|---|---|---|---|---|
| CN1 | Slurred speech | 0.975 | 0.094 | 0.525 | 0.592 | | | | | | |
| CN2 | Hypersensitivity to sound/light | -0.9 | 0.081 | -0.532 | 0.241 | -0.018 | | | | | |
| CN3 | Hypersensitivity to tastes /smells | 0.196 | 0.078 | -1.659 | 0.010 | -0.214 | -0.102 | | | | |
| CN4 | Tinnitus | -0.336 | 0.068 | 1.843 | 0.520 | -0.270 | -0.267 | -0.209 | | | |
| CN5 | Visual difficulty/ double vision | 0.388 | 0.083 | -0.244 | 0.014 | -0.071 | -0.23 | -0.176 | -0.286 | | |
| CN6 | Dry eyes | -0.322 | 0.074 | -0.306 | 0.448 | -0.204 | -0.293 | -0.132 | -0.286 | -0.087 | |
| | | | | | Average residual correlation | -0.190 | | | | | |
| | | | | | LD criterion (ave + 0.2) | 0.010 | | | | | |

### Immune System Domain Individual Item Fit Table

| Item Code | Statement | Location | SE | Fit Residual | Chi-Square P | Local Dependency | | | |
|---|---|---|---|---|---|---|---|---|---|
| | | | | | | IM1 | IM2 | IM3 | IM4 |
| IM1 | Sore throat/ voice | 0.129 | 0.096 | 0.493 | 0.897 | | | | |
| IM2 | Swollen/tender lymph nodes | 0.297 | 0.092 | -0.614 | 0.247 | -0.171 | | | |
| IM3 | Pyrexia | 0.072 | 0.095 | 0.013 | 0.196 | -0.236 | -0.361 | | |
| IM4 | Allergic reactions | -0.498 | 0.085 | 1.29 | 0.445 | -0.496 | -0.356 | -0.331 | |
| | | | | | Average residual correlation | -0.325 | | | |
| | | | | | LD criterion (ave + 0.2) | -0.125 | | | |
