## Supplementary File 2 Individ Item Fit Subscales for "Development and psychometric evaluation of The Index of Myalgic Encephalomyelitis Symptoms (TIMES) Part I: Rasch Analysis and Content Validity"

**Supplementary File 2 – Rasch Analysis Final Individual Item Fit Tables for TIMES Subscales and Total score, using domain-level super-items**

Any highlighted (coloured) values fall outside of fit criteria stated in Methods section

### Dysautonomia Subscale Individual Item Fit Table (Domain-level super-items)

| | | | | | | Local Dependency | | | | |
|---|---|---|---|---|---|---|---|---|---|---|
| Item Code | Statement | Location | SE | Fit Residual | Chi-Square P | CNDom | GIDom | CRDom | SLDom | ISDom |
| CNDom | Cranial Nerves | 0.08 | 0.026 | -1.515 | 0.537 | | | | | |
| GIDom | Gastro-Intestinal | 0.254 | 0.024 | 0.063 | 0.348 | -0.257 | | | | |
| CRDom | Cardio-Respiratory | -0.052 | 0.02 | -1.212 | 0.866 | -0.161 | -0.315 | | | |
| SLDom | Sleep | -0.496 | 0.024 | 3.234 | 0.109 | -0.304 | -0.214 | -0.464 | | |
| ISDom | Immune System | 0.213 | 0.031 | -0.465 | 0.405 | -0.079 | -0.219 | -0.152 | -0.244 | |
| | | | | Average residual correlation | | -0.241 | | | | |
| | | | | LD criterion (ave + 0.2) | | -0.041 | | | | |

### Neurology Subscale Individual Item Fit Table (Domain-level super-items)

| | | | | | | Local Dependency | | |
|---|---|---|---|---|---|---|---|---|
| Item | Statement | Location | SE | FitResid | Prob | CODom | PADom | MSDom |
| CODom | Cognitive | -0.334 | 0.017 | 2.539 | 0.056 | | | |
| PADom | Pain | 0.241 | 0.023 | -0.082 | 0.061 | -0.775 | | |
| MSDom | Motor-Sensory | 0.093 | 0.021 | -1.069 | 0.081 | -0.801 | 0.264 | |
| | | | | Average residual correlation | | -0.437 | | |
| | | | | LD criterion (ave + 0.2) | | -0.237 | | |

### TIMES-Total Full Scale Individual Item Fit Table (Domain-level super-items)

| | | | | | | Local Dependency | | | | | | | | |
|---|---|---|---|---|---|---|---|---|---|---|---|---|---|---|
| Item | Statement | Location | SE | FitResid | Prob | FADom | CODom | PADom | MSDom | CNDom | GIDom | CRDom | SLDom | ISDom |
| FADom | Fatigue | -1.564 | 0.03 | 1.617 | 0.963 | | | | | | | | | |
| CODom | Cognitive | -0.301 | 0.017 | 5.198 | 0.000 | 0.163 | | | | | | | | |
| PADom | Pain | 0.309 | 0.024 | -1.882 | 0.087 | -0.186 | -0.428 | | | | | | | |
| MSDom | Motor-Sensory | 0.145 | 0.022 | -2.887 | 0.029 | -0.280 | -0.425 | 0.325 | | | | | | |
| CNDom | Cranial Nerves | 0.358 | 0.025 | -2.556 | 0.059 | -0.275 | -0.189 | 0.070 | 0.013 | | | | | |
| GIDom | Gastro-Intestinal | 0.56 | 0.024 | 1.01 | 0.050 | -0.216 | -0.185 | -0.193 | -0.127 | -0.080 | | | | |
| CRDom | Cardio-Respiratory | 0.232 | 0.02 | -0.143 | 0.478 | -0.138 | -0.324 | -0.111 | -0.066 | -0.001 | -0.032 | | | |
| SLDom | Sleep | -0.215 | 0.024 | 1.404 | 0.257 | 0.043 | -0.045 | -0.081 | -0.090 | -0.274 | -0.105 | -0.330 | | |
| ISDom | Immune System | 0.476 | 0.03 | -0.505 | 0.137 | -0.210 | -0.364 | 0.057 | 0.028 | 0.060 | -0.001 | 0.077 | -0.150 | |
| | | | | | Average residual correlation | -0.113 | | | | | | | | |
| | | | | | LD criterion (ave + 0.2) | 0.087 | | | | | | | | |
