## Supplementary file 3. Rasch ordinal to interval conversion tables for "Development and psychometric evaluation of The Index of Myalgic Encephalomyelitis Symptoms (TIMES) Part I: Rasch Analysis and Content Validity"

**TIMES2 Full Scale Total Score** (0-174) to 0-100 Linear Conversion Chart

| Raw Score | 0-100 Conversion |
|---|---|
| **0** | 0 |
| **1** | 14.58 |
| **2** | 23.26 |
| **3** | 28.41 |
| **4** | 31.85 |
| **5** | 34.36 |
| **6** | 36.32 |
| **7** | 37.91 |
| **8** | 39.24 |
| **9** | 40.4 |
| **10** | 41.41 |
| **11** | 42.31 |
| **12** | 43.12 |
| **13** | 43.86 |
| **14** | 44.54 |
| **15** | 45.17 |
| **16** | 45.76 |
| **17** | 46.31 |
| **18** | 46.82 |
| **19** | 47.32 |
| **20** | 47.78 |
| **21** | 48.23 |
| **22** | 48.66 |
| **23** | 49.07 |
| **24** | 49.45 |
| **25** | 49.83 |
| **26** | 50.19 |
| **27** | 50.54 |
| **28** | 50.88 |
| **29** | 51.2 |
| **30** | 51.53 |
| **31** | 51.83 |
| **32** | 52.14 |
| **33** | 52.42 |
| **34** | 52.7 |
| **35** | 52.98 |
| **36** | 53.25 |
| **37** | 53.52 |
| **38** | 53.78 |
| **39** | 54.03 |
| **40** | 54.27 |
| **41** | 54.51 |
| **42** | 54.75 |
| **43** | 54.98 |
| **44** | 55.21 |
| **45** | 55.43 |
| **46** | 55.65 |
| **47** | 55.86 |
| **48** | 56.08 |
| **49** | 56.28 |
| **50** | 56.49 |
| **51** | 56.69 |
| **52** | 56.89 |
| **53** | 57.08 |
| **54** | 57.27 |
| **55** | 57.47 |
| **56** | 57.66 |
| **57** | 57.84 |
| **58** | 58.03 |
| **59** | 58.2 |
| **60** | 58.38 |
| **61** | 58.56 |
| **62** | 58.73 |
| **63** | 58.91 |
| **64** | 59.08 |
| **65** | 59.25 |
| **66** | 59.41 |
| **67** | 59.58 |
| **68** | 59.74 |
| **69** | 59.91 |
| **70** | 60.07 |
| **71** | 60.23 |
| **72** | 60.39 |
| **73** | 60.55 |
| **74** | 60.7 |
| **75** | 60.86 |
| **76** | 61.02 |
| **77** | 61.17 |
| **78** | 61.32 |
| **79** | 61.48 |
| **80** | 61.63 |
| **81** | 61.78 |
| **82** | 61.93 |
| **83** | 62.08 |
| **84** | 62.23 |
| **85** | 62.38 |
| **86** | 62.53 |
| **87** | 62.67 |
| **88** | 62.82 |
| **89** | 62.96 |
| **90** | 63.11 |
| **91** | 63.26 |
| **92** | 63.41 |
| **93** | 63.55 |
| **94** | 63.7 |
| **95** | 63.84 |
| **96** | 63.99 |
| **97** | 64.13 |
| **98** | 64.28 |
| **99** | 64.43 |
| **100** | 64.57 |
| **101** | 64.72 |
| **102** | 64.86 |
| **103** | 65.01 |
| **104** | 65.15 |
| **105** | 65.3 |
| **106** | 65.45 |
| **107** | 65.6 |
| **108** | 65.74 |
| **109** | 65.89 |
| **110** | 66.05 |
| **111** | 66.2 |
| **112** | 66.35 |
| **113** | 66.5 |
| **114** | 66.66 |
| **115** | 66.81 |
| **116** | 66.97 |
| **117** | 67.12 |
| **118** | 67.28 |
| **119** | 67.45 |
| **120** | 67.6 |
| **121** | 67.77 |
| **122** | 67.93 |
| **123** | 68.1 |
| **124** | 68.27 |
| **125** | 68.44 |
| **126** | 68.61 |
| **127** | 68.78 |
| **128** | 68.95 |
| **129** | 69.14 |
| **130** | 69.32 |
| **131** | 69.5 |
| **132** | 69.69 |
| **133** | 69.88 |
| **134** | 70.07 |
| **135** | 70.27 |
| **136** | 70.47 |
| **137** | 70.67 |
| **138** | 70.88 |
| **139** | 71.09 |
| **140** | 71.31 |
| **141** | 71.54 |
| **142** | 71.76 |
| **143** | 71.99 |
| **144** | 72.23 |
| **145** | 72.48 |
| **146** | 72.73 |
| **147** | 72.98 |
| **148** | 73.26 |
| **149** | 73.53 |
| **150** | 73.81 |
| **151** | 74.11 |
| **152** | 74.42 |
| **153** | 74.73 |
| **154** | 75.06 |
| **155** | 75.4 |
| **156** | 75.77 |
| **157** | 76.14 |
| **158** | 76.54 |
| **159** | 76.97 |
| **160** | 77.42 |
| **161** | 77.9 |
| **162** | 78.42 |
| **163** | 78.98 |
| **164** | 79.59 |
| **165** | 80.26 |
| **166** | 81.01 |
| **167** | 81.86 |
| **168** | 82.85 |
| **169** | 84.01 |
| **170** | 85.42 |
| **171** | 87.25 |
| **172** | 89.76 |
| **173** | 93.73 |
| **174** | 100 |

**Dysautonomia Subscale Score** (0-96 to 0-100 Linear Conversion Chart

| Raw Score | 0-100 Conversion |
|---|---|
| **0** | 0 |
| **1** | 9.04 |
| **2** | 15 |
| **3** | 18.92 |
| **4** | 21.87 |
| **5** | 24.25 |
| **6** | 26.24 |
| **7** | 27.96 |
| **8** | 29.5 |
| **9** | 30.89 |
| **10** | 32.15 |
| **11** | 33.31 |
| **12** | 34.38 |
| **13** | 35.39 |
| **14** | 36.35 |
| **15** | 37.24 |
| **16** | 38.1 |
| **17** | 38.9 |
| **18** | 39.68 |
| **19** | 40.44 |
| **20** | 41.16 |
| **21** | 41.87 |
| **22** | 42.53 |
| **23** | 43.19 |
| **24** | 43.82 |
| **25** | 44.42 |
| **26** | 45.03 |
| **27** | 45.61 |
| **28** | 46.18 |
| **29** | 46.72 |
| **30** | 47.26 |
| **31** | 47.78 |
| **32** | 48.29 |

| Raw Score | 0-100 Conversion |
|---|---|
| **33** | 48.8 |
| **34** | 49.28 |
| **35** | 49.75 |
| **36** | 50.23 |
| **37** | 50.69 |
| **38** | 51.14 |
| **39** | 51.58 |
| **40** | 52.02 |
| **41** | 52.44 |
| **42** | 52.87 |
| **43** | 53.28 |
| **44** | 53.69 |
| **45** | 54.09 |
| **46** | 54.49 |
| **47** | 54.9 |
| **48** | 55.29 |
| **49** | 55.68 |
| **50** | 56.06 |
| **51** | 56.45 |
| **52** | 56.82 |
| **53** | 57.2 |
| **54** | 57.59 |
| **55** | 57.97 |
| **56** | 58.35 |
| **57** | 58.73 |
| **58** | 59.11 |
| **59** | 59.5 |
| **60** | 59.87 |
| **61** | 60.26 |
| **62** | 60.66 |
| **63** | 61.06 |
| **64** | 61.45 |
| **65** | 61.85 |

| Raw Score | 0-100 Conversion |
|---|---|
| **66** | 62.27 |
| **67** | 62.68 |
| **68** | 63.1 |
| **69** | 63.53 |
| **70** | 63.96 |
| **71** | 64.4 |
| **72** | 64.85 |
| **73** | 65.32 |
| **74** | 65.78 |
| **75** | 66.28 |
| **76** | 66.77 |
| **77** | 67.28 |
| **78** | 67.81 |
| **79** | 68.36 |
| **80** | 68.94 |
| **81** | 69.53 |
| **82** | 70.14 |
| **83** | 70.8 |
| **84** | 71.51 |
| **85** | 72.25 |
| **86** | 73.06 |
| **87** | 73.94 |
| **88** | 74.91 |
| **89** | 76.01 |
| **90** | 77.27 |
| **91** | 78.74 |
| **92** | 80.57 |
| **93** | 82.91 |
| **94** | 86.19 |
| **95** | 91.48 |
| **96** | 100 |

**Neurological Subscale Score** (0-66) to 0-100 Linear Conversion Chart

| Raw Score | 0-100 Conversion |
|---|---|
| **0** | 0 |
| **1** | 9.48 |
| **2** | 15.66 |
| **3** | 19.68 |
| **4** | 22.71 |
| **5** | 25.18 |
| **6** | 27.31 |
| **7** | 29.18 |
| **8** | 30.86 |
| **9** | 32.4 |
| **10** | 33.82 |
| **11** | 35.14 |
| **12** | 36.37 |
| **13** | 37.52 |
| **14** | 38.62 |
| **15** | 39.63 |
| **16** | 40.62 |
| **17** | 41.54 |
| **18** | 42.42 |
| **19** | 43.26 |
| **20** | 44.06 |
| **21** | 44.83 |
| **22** | 45.57 |
| **23** | 46.28 |
| **24** | 46.97 |
| **25** | 47.63 |
| **26** | 48.28 |
| **27** | 48.89 |
| **28** | 49.51 |
| **29** | 50.11 |
| **30** | 50.69 |
| **31** | 51.28 |
| **32** | 51.86 |
| **33** | 52.43 |

| Raw Score | 0-100 Conversion |
|---|---|
| **34** | 53 |
| **35** | 53.55 |
| **36** | 54.12 |
| **37** | 54.69 |
| **38** | 55.26 |
| **39** | 55.83 |
| **40** | 56.42 |
| **41** | 56.98 |
| **42** | 57.58 |
| **43** | 58.18 |
| **44** | 58.78 |
| **45** | 59.42 |
| **46** | 60.05 |
| **47** | 60.69 |
| **48** | 61.37 |
| **49** | 62.06 |
| **50** | 62.78 |
| **51** | 63.54 |
| **52** | 64.32 |
| **53** | 65.15 |
| **54** | 66.03 |
| **55** | 66.97 |
| **56** | 67.98 |
| **57** | 69.09 |
| **58** | 70.32 |
| **59** | 71.69 |
| **60** | 73.28 |
| **61** | 75.14 |
| **62** | 77.4 |
| **63** | 80.26 |
| **64** | 84.18 |
| **65** | 90.34 |
| **66** | 100 |

Raw Score to 0-100 Linear Score Conversion Charts for **Cardio-Respiratory**, **Cognitive**, and **Gastro-Intestinal** Domains

| Cardio-Respiratory Domain (0-27) | |
|---|---|
| **Raw Score** | **0-100 Conversion** |
| **0** | 0 |
| **1** | 9.48 |
| **2** | 15.66 |
| **3** | 19.68 |
| **4** | 22.71 |
| **5** | 25.18 |
| **6** | 27.31 |
| **7** | 29.18 |
| **8** | 30.86 |
| **9** | 32.4 |
| **10** | 33.82 |
| **11** | 35.14 |
| **12** | 36.37 |
| **13** | 37.52 |
| **14** | 38.62 |
| **15** | 39.63 |
| **16** | 40.62 |
| **17** | 41.54 |
| **18** | 42.42 |
| **19** | 43.26 |
| **20** | 44.06 |
| **21** | 44.83 |
| **22** | 45.57 |
| **23** | 46.28 |
| **24** | 46.97 |
| **25** | 47.63 |
| **26** | 48.28 |
| **27** | 48.89 |

| Cognitive Domain (0-27) | |
|---|---|
| **Raw Score** | **0-100 Conversion** |
| **0** | 53 |
| **1** | 53.55 |
| **2** | 54.12 |
| **3** | 54.69 |
| **4** | 55.26 |
| **5** | 55.83 |
| **6** | 56.42 |
| **7** | 56.98 |
| **8** | 57.58 |
| **9** | 58.18 |
| **10** | 58.78 |
| **11** | 59.42 |
| **12** | 60.05 |
| **13** | 60.69 |
| **14** | 61.37 |
| **15** | 62.06 |
| **16** | 62.78 |
| **17** | 63.54 |
| **18** | 64.32 |
| **19** | 65.15 |
| **20** | 66.03 |
| **21** | 66.97 |
| **22** | 67.98 |
| **23** | 69.09 |
| **24** | 70.32 |
| **25** | 71.69 |
| **26** | 73.28 |
| **27** | 75.14 |

| Gastro-Intestinal Domain (0-21) | |
|---|---|
| **Raw Score** | **0-100 Conversion** |
| **0** | 0 |
| **1** | 9.48 |
| **2** | 15.66 |
| **3** | 19.68 |
| **4** | 22.71 |
| **5** | 25.18 |
| **6** | 27.31 |
| **7** | 29.18 |
| **8** | 30.86 |
| **9** | 32.4 |
| **10** | 33.82 |
| **11** | 35.14 |
| **12** | 36.37 |
| **13** | 37.52 |
| **14** | 38.62 |
| **15** | 39.63 |
| **16** | 40.62 |
| **17** | 41.54 |
| **18** | 42.42 |
| **19** | 43.26 |
| **20** | 44.06 |
| **21** | 44.83 |

Raw Score to 0-100 Linear Score Conversion Charts for **Motor-Sensory**, **Cranial Nerves**, and **Pain** Domains

| Motor-Sensory Domain (0-21) | |
|---|---|
| Raw Score | 0-100 Conversion |
| 0 | 53 |
| 1 | 53.55 |
| 2 | 54.12 |
| 3 | 54.69 |
| 4 | 55.26 |
| 5 | 55.83 |
| 6 | 56.42 |
| 7 | 56.98 |
| 8 | 57.58 |
| 9 | 58.18 |
| 10 | 58.78 |
| 11 | 59.42 |
| 12 | 60.05 |
| 13 | 60.69 |
| 14 | 61.37 |
| 15 | 62.06 |
| 16 | 62.78 |
| 17 | 63.54 |
| 18 | 64.32 |
| 19 | 65.15 |
| 20 | 66.03 |
| 21 | 66.97 |

| Cranial Nerves Domain (0-18) | |
|---|---|
| Raw Score | 0-100 Conversion |
| 0 | 0 |
| 1 | 9.48 |
| 2 | 15.66 |
| 3 | 19.68 |
| 4 | 22.71 |
| 5 | 25.18 |
| 6 | 27.31 |
| 7 | 29.18 |
| 8 | 30.86 |
| 9 | 32.4 |
| 10 | 33.82 |
| 11 | 35.14 |
| 12 | 36.37 |
| 13 | 37.52 |
| 14 | 38.62 |
| 15 | 39.63 |
| 16 | 40.62 |
| 17 | 41.54 |
| 18 | 42.42 |

| Pain Domain (0-18) | |
|---|---|
| Raw Score | 0-100 Conversion |
| 0 | 53 |
| 1 | 53.55 |
| 2 | 54.12 |
| 3 | 54.69 |
| 4 | 55.26 |
| 5 | 55.83 |
| 6 | 56.42 |
| 7 | 56.98 |
| 8 | 57.58 |
| 9 | 58.18 |
| 10 | 58.78 |
| 11 | 59.42 |
| 12 | 60.05 |
| 13 | 60.69 |
| 14 | 61.37 |
| 15 | 62.06 |
| 16 | 62.78 |
| 17 | 63.54 |
| 18 | 64.32 |

Raw Score to 0-100 Linear Score Conversion Charts for **Sleep**, **Immune System**, and **Fatigue** Domains

| Sleep Domain (0-18) | |
|---|---|
| **Raw Score** | **0-100 Conversion** |
| **0** | 0 |
| **1** | 9.48 |
| **2** | 15.66 |
| **3** | 19.68 |
| **4** | 22.71 |
| **5** | 25.18 |
| **6** | 27.31 |
| **7** | 29.18 |
| **8** | 30.86 |
| **9** | 32.4 |
| **10** | 33.82 |
| **11** | 35.14 |
| **12** | 36.37 |
| **13** | 37.52 |
| **14** | 38.62 |
| **15** | 39.63 |
| **16** | 40.62 |
| **17** | 41.54 |
| **18** | 42.42 |

| Immune System Domain (0-12) | |
|---|---|
| **Raw Score** | **0-100 Conversion** |
| **0** | 53 |
| **1** | 53.55 |
| **2** | 54.12 |
| **3** | 54.69 |
| **4** | 55.26 |
| **5** | 55.83 |
| **6** | 56.42 |
| **7** | 56.98 |
| **8** | 57.58 |
| **9** | 58.18 |
| **10** | 58.78 |
| **11** | 59.42 |
| **12** | 60.05 |

| Fatigue Domain (0-12) | |
|---|---|
| **Raw Score** | **0-100 Conversion** |
| **0** | 0 |
| **1** | 9.48 |
| **2** | 15.66 |
| **3** | 19.68 |
| **4** | 22.71 |
| **5** | 25.18 |
| **6** | 27.31 |
| **7** | 29.18 |
| **8** | 30.86 |
| **9** | 32.4 |
| **10** | 33.82 |
| **11** | 35.14 |
| **12** | 36.37 |
