## Supplementary file 4 TIMES final version for "Development and psychometric evaluation of The Index of Myalgic Encephalomyelitis Symptoms (TIMES) Part I: Rasch Analysis and Content Validity"

### The Index of ME Symptoms (TIMES)

The aim of this questionnaire is to assess your symptoms. It has been developed by people with ME/CFS and clinicians working in specialist ME/CFS services and is intended to be used with the other assessment tools in the ME Association's Clinical Assessment Toolkit.

It asks about which symptoms you are experiencing, and how troublesome they are. This can be very variable, so the questions aim to take a snapshot of how, overall, each symptom is affecting you at present (i.e. on an average day over the last month), rather than recording the whole history, comparing you to other people, or how you were before you became ill. This can help with making a diagnosis and help you and other people understand the impact of your symptoms and start discussions about how to manage them.

There are questions about 58 symptoms, arranged in several sections so the questionnaire is quite long. This is because it is important to cover everything. However, it uses quick and simple multiple-choice answers and there also opportunity to add further detail if you wish. It takes about 15 minutes to complete in one go, but you can take as long as you want to complete it. Your answers will be saved automatically. If you need help from another person, or another person to complete the TIMES on your behalf, that is fine. If you would prefer a paper copy or complete the survey by phone, or if you have any other questions or need further adjustments to make it easier for you to complete this questionnaire, please contact the person who sent this form.

NB– each section (ie fatigue; neurological symptoms; dysautonomia) can be used separately as 'stand alone' assessments, as can the sub-scales.

#### Section 1. Fatigue

| **Over the last month, how often have you experienced this symptom?** | I do not have this symptom (score 0) | Some of the time (score 1) | Most of the time (score 2) | All the time (score 3) |
|---|---|---|---|---|
| Physical exhaustion after previously undemanding activity | | | | |
| Loss of physical strength, or stamina during/ after a previously undemanding activity | | | | |
| Cognitive/ mental exhaustion (also known as 'brain fog') after previously undemanding activity | | | | |

| |
|---|
| Post exertional malaise (PEM). *PEM describes a worsening of symptoms after seemingly trivial or undemanding activity of any description. It is often referred to as 'a crash'. Onset may be delayed and it can be long lasting.* |
| Total fatigue score (add the scores for the questions above) |

#### SECTION 2: NEUROLOGICAL SYMPTOMS

**SECTION 2a COGNITIVE SYMPTOMS** NB. If you are unable to do any of the activities listed, then answer 'all the time'

| **Over the last month, how often have you experienced this symptom?** | I do not have this symptom (score 0) | Some of the time (score 1) | Most of the time (score 2) | All the time (score 3) |
|---|---|---|---|---|
| Memory and/or concentration problems. (e.g. forgetfulness; difficulty concentrating; being easily distracted; losing your train of thought or track of conversations) | | | | |
| Slowness of thoughts and/ or reactions | | | | |
| Difficulty starting and/ or finishing tasks. | | | | |
| Difficulty making decisions and problem-solving ('working things out'). | | | | |
| Difficulty getting organised. | | | | |
| Difficulty multi-tasking (doing more than one thing at once e.g. walking and talking) | | | | |
| Difficulty taking in or retaining information. | | | | |
| Communication difficulties: Difficulty finding the right words; getting words (or numbers) jumbled up. | | | | |
| Difficulty reading and/ or writing. | | | | |

| Total cognitive score (add the scores for the questions above) |
|---|

**SECTION 2b PAIN**

| **Over the last month, how troublesome has this symptom been?** | I do not have this symptom (score 0) | Mild-Moderate. Interfering with some activities (score 1) | Severe. Interfering with most/all activities (score 2) | Very severe. Unable to carry out activities (score 3) |
|---|---|---|---|---|
| Musculo-skeletal (muscle, joint, and/or bone) pain | | | | |
| Jaw Pain | | | | |
| Eye pain | | | | |
| Nerve pain or neuralgia | | | | |
| Headaches and/or migraines. *A migraine is typically a severe headache with a throbbing pain on one side of the head. There may be warning signs beforehand such as dizziness; visual changes, numbness or pins and needles.* | | | | |
| Allodynia, i.e. Pain and /or tenderness from sensations others would not find painful e.g. touch, sound, visual stimuli, temperature | | | | |
| Total pain score (add the scores for the questions above) | | | | |

**SECTION 2c. MOTOR-SENSORY SYSTEM SYMPTOMS**

| **Over the last month, how troublesome has this symptom been?** | I do not have this symptom (score 0) | Mild-Moderate. Interfering with some | Severe. Interfering with most/all | Very severe. Unable to carry out |
|---|---|---|---|---|

| | | activities (score 1) | activities (score 2) | activities (score 3) |
|---|---|---|---|---|
| Muscle tightness | | | | |
| Muscle cramp or twitches, jerks and/or spasms | | | | |
| Tremors: shakiness or wobbliness in the limbs, head or trunk | | | | |
| Slow and/or weak movement | | | | |
| Clumsiness and/or balance problems (e.g. dropping or knocking things over; tripping or slipping; knees giving way, or catching your toes) | | | | |
| Increased sensitivity to touch or pressure | | | | |
| Numbness or altered sensation (e.g. tingling, stabbing, burning, feeling something is crawling/ running over your skin) | | | | |
| Total motor-sensory score (add the scores for the questions above) | | | | |

#### SECTION 3: DYSAUTONOMIA

##### SECTION 3a SLEEP SYMPTOMS

| **Over the last month, how troublesome has this symptom been?** | I do not have this symptom (score 0) | Mild-Moderate. (score 1) | Severe. (score 2) | Very severe. (score 3) |
|---|---|---|---|---|
| Change to sleep pattern (e.g. changing from an early bird to a night owl or having 'day-night reversal'). | | | | |

| |
|---|
| Needing to nap or sleep during the day |
| Difficulty falling asleep when you want to sleep (during the day or night) |
| Difficulty staying asleep (also known as sleep maintenance) i.e. waking frequently during the night and/ or in the early hours of the morning |
| Taking a long time to 'come to' on waking (i.e. it may take some time to become aware and feel able to move/ function). |
| Feeling exhausted, flu-like or stiff on waking |
| Total sleep score (add the scores for the questions above) |

**SECTION 3b. CARDIO-RESPIRATORY SYMPTOMS**

| **Over the last month, how troublesome has this symptom been?** | I do not have this symptom (score 0) | Mild-Moderate. Interfering with some activities (score 1) | Severe. Interfering with most/all activities (score 2) | Very severe. Unable to carry out activities (score 3) |
|---|---|---|---|---|
| Increased sensitivity / intolerance to temperature (e.g. hot food or drinks; temperature of the room; hot or cold weather). *This may involve sweating, hot flushes, chills, or feeling hot or cold in temperatures that others would fine unremarkable* | | | | |
| Dizziness / vertigo/ light-headedness | | | | |
| Palpitations: fast or irregular heartbeats during/ after | | | | |

| |
|---|
| previously undemanding activity, or at rest |
| Chest pain at rest or during/after previously undemanding activity |
| Shortness of breath or trouble catching your breath. At rest, or during /after previously undemanding activity |
| Poor circulation: Cold hands and/or feet which is not caused by the temperature of the surroundings and/or inability to warm up promptly after becoming cold |
| Orthostatic intolerance: *Increase in symptoms (e.g. dizziness, blurred vision, palpitations, breathlessness, headache, nausea) when changing to a more upright position. (e.g. standing up) or sitting or standing for long periods. Symptoms may ease when you sit or lie with their feet up. Or you may be able to do more cognitive activity (e.g. reading, talking, or desk work) when lying down rather than when sitting or standing). NB. If you are unable to sit or stand, answer 'very severe'.* |
| Swollen or discoloured (usually pink or purple). extremities (hands /fingers or feet/toes) if in upright position (sitting or standing) for a long time. *NB. If you are unable to sit or stand, answer 'very severe'.* |
| Abnormal sweating (e.g. night sweats/ hot flushes or chills) |

| |
|---|
| Total cardio-respiratory score (add the scores for the questions above) |

**SECTION 3c. CRANIAL NERVES**

| **Over the last month, how troublesome has this symptom been?** | I do not have this symptom (score 0) | Mild-Moderate. (score 1) | Severe. (score 2) | Very severe. (score 3) |
|---|---|---|---|---|
| Slow or slurred speech | | | | |
| Increased sensitivity to sounds and/or sensitivity to light/ moving images | | | | |
| Increased or decreased sensitivity to tastes and/or smells including 'phantom' smells which aren't really there. | | | | |
| Tinnitus or 'ringing in the ears' | | | | |
| Blurred/ double vision/ difficulty focussing | | | | |
| Dry eyes or mouth | | | | |
| Total cranial nerves score (add the scores for the questions above) | | | | |

**SECTION 3d. GASTRO-INTESTINAL SYMPTOMS**

| **Over the last month, how troublesome has this symptom been?** | I do not have this symptom (score 0) | Mild-Moderate. (score 1) | Severe. (score 2) | Very severe. (score 3) |
|---|---|---|---|---|
| Nausea and/or vomiting (feeling or being sick) | | | | |
| Abdominal pain and/or bloating | | | | |
| Excessive flatulence (farting a lot) | | | | |
| Changes in bowel habit: diarrhoea, constipation, urgency and /or frequency of defecation (having a poo) | | | | |

| |
|---|
| Change of appetite: increase or decrease). NB. If you are unable to eat, answer 'very severe' |
| Difficulty eating and drinking (e.g. biting, chewing, swallowing). NB. If you are unable to eat, answer 'very severe' |
| Being too tired to eat. NB. If you are unable to eat, answer 'very severe' |
| Total gastro-intestinal score (add the scores for the questions above) |

**SECTION 3E. IMMUNE SYSTEM SYMPTOMS**

| **Over the last month, how troublesome has this symptom been?** | I do not have this symptom (score 0) | Mild-Moderate. (score 1) | Severe (score 2) | Very severe. (score 3) |
|---|---|---|---|---|
| Sore throat or hoarse voice | | | | |
| Tender lymph nodes in the armpits, groin and/or neck | | | | |
| Pyrexia or fever: *Feeling like you have a raised temperature with hot sweats and/or chills, although it may be normal when measured* | | | | |
| Allergic reactions *(e.g. runny eyes, stuffy nose, cough, abdominal pain, feeling sick, flushed/blotchy/itchy skin, rashes, headache, wheezing or breathlessness) to smells, tastes, foods, medications, plants, or chemicals.* | | | | |
| Total immune system score (add the scores for the questions above) | | | | |

**FINAL SECTION**

Is there anything else about your symptoms you would like us to know? (free text)

### The Index of ME Symptoms (TIMES) Summary Report

This report summarises your responses to The Index of ME Symptoms questionnaire (TIMES) which evaluates your symptoms,

| | |
|---|---|
| Name: | NHS Number |
| Date: | No of assessment |

**Here is your symptom profile**

| | **Score** | **Interpretation** |
|---|---|---|
| **1. Fatigue** | | 0-3 = mild; 4-6= moderate; 6-8= severe; 9-12= very severe |
| 2a. Cognition | | 0-7 = mild; 8-13= moderate; 14-20= severe; 21-28= very severe |
| 2b. Pain | | 0-5=mild; 6-9= moderate; 10-13= severe; 14-18=very severe |
| 2c. Motor-sensory symptoms | | 0-5= mild; 6-10 = moderate; 11-15= severe; 16-21 = very severe |
| 2. **Neurological Symptoms** (add up the scores for sections 2a-2c) | | 0-16 = mild; 17-33 = moderate; 34-49 = severe; 50-66 = very severe) |
| 3a. Sleep | | 0-5=mild; 6-9= moderate; 10-13= severe; 14-18=very severe |
| 3b. Cardio-respiratory symptoms | | 0-7= mild; 8-14 = moderate; 15-21 severe; 22-27 = very severe |
| 3c. Cranial nerves | | 0-5=mild; 6-9= moderate; 10-13= severe; 14-18=very severe |
| 3d. Gastro-intestinal symptoms | | 0-5= mild; 6-10 = moderate; 11-15= severe; 16-21 = very severe |
| 3e. Immune system | | 0-3 = mild; 4-6 = moderate; 7-9 = severe; 10-12 = very severe |
| 3. **Dysautonomia** (add up the scores for sections 3a-3e) | | 0-24 = mild; 26-48 = moderate; 49-72 = severe; 73-96 = very severe |
| **4. Total TIMES Score** (add up the scores for fatigue, | | 0-44 = mild; 45-87 = moderate;88-131 = severe; 132-174 = very severe |

| |
|---|
| neurological and dysautonomia scales) |
| Any other comments re: your symptoms |
